# Clinical Evaluation of a Combo Rapid Antigen Test QuickNavi-Flu+COVID19 Ag for Simultaneous Detection of SARS-CoV-2 and Influenza Viruses

**DOI:** 10.1101/2021.12.05.21267215

**Authors:** Yuto Takeuchi, Yusaku Akashi, Yoshihiko Kiyasu, Norihiko Terada, Yoko Kurihara, Daisuke Kato, Takashi Miyazawa, Shino Muramatsu, Yuki Shinohara, Atsuo Ueda, Shigeyuki Notake, Koji Nakamura, Hiromichi Suzuki

## Abstract

**Introduction:** Since respiratory sample collection is an uncomfortable experience, simultaneous detection of pathogens with a single swab is preferable. We prospectively evaluated the clinical performance of a newly developed antigen test QuickNavi-Flu+COVID19 Ag (Denka Co., Ltd., Tokyo, Japan) which can detect severe acute respiratory syndrome coronavirus 2 (SARS-CoV-2) and influenza viruses at the same time with a single testing device.

**Methods:** Included were those who were suspected of contracting coronavirus disease 2019 (COVID-19) and referred to a PCR center at Ibaraki prefecture in Japan, between August 2, 2021 to September 13, 2021, when the L452R mutant strains of SARS-CoV-2 were prevalent. Additional nasopharyngeal samples and anterior nasal samples were obtained for the antigen test and were compared with a reference reverse transcription PCR (RT-PCR) using nasopharyngeal samples.

**Results:** In total, 1510 nasopharyngeal samples and 862 anterior nasal samples were evaluated. For SARS-CoV-2 detection in nasopharyngeal samples, the sensitivity and specificity of the antigen test were 80.9% and 99.8%, respectively. The sensitivity and specificity using anterior nasal samples were 67.8% and 100%, respectively. In symptomatic cases, the sensitivities increased to 88.3% with nasopharyngeal samples and 73.7% with anterior nasal samples. There were three cases of discrepant results between the antigen test and the real-time RT-PCR. All of them were positive with the antigen test but negative with the real-time RT-PCR in SARS-CoV-2 detection. During the study period, influenza viruses were not detected.

**Conclusion:** A combo kit, QuickNavi-Flu+COVID19 Ag, showed an acceptable sensitivity and sufficient specificity for SARS-CoV-2 detection, especially using nasopharyngeal sample collected from symptomatic patients.

---

The severe acute respiratory syndrome coronavirus 2 (SARS-CoV-2) pandemic is still continuing worldwide and remains a major health problem [1]. In addition, although there is a concern about the simultaneous outbreak of SARS-CoV-2 and influenza viruses in winter season, it can be difficult to distinguish coronavirus disease 2019 (COVID-19) from influenza based on clinical symptoms [2].

Antigen testing is simpler and faster than PCR testing and therefore is expected to play a role in COVID-19 infection control [3]. In general, it is necessary to collect separate samples for SARS-CoV-2 and for influenza viruses to perform two different antigen testing, though the multiple collections of respiratory samples are accompanied by discomfort. The newly developed combo rapid antigen test QuickNavi-Flu+COVID19 Ag (Denka Co., Ltd., Tokyo, Japan) consists of the main parts of QuickNavi-COVID19 Ag (Denka Co., Ltd., Tokyo, Japan) and QuickNavi-Flu2 (Denka Co., Ltd., Tokyo, Japan) that are already in market. QuickNavi-Flu+COVID19 Ag can detect SARS-CoV-2 and influenza viruses simultaneously using nasopharyngeal or anterior nasal specimens, and the read time has been shortened compared with QuickNavi-COVID19 Ag [4]. Although QuickNavi-Flu+COVID19 Ag is commercially available in Japan, its usefulness in clinical practice has not yet been fully assessed.

In this study, we prospectively evaluated the clinical performance of the combo kit QuickNavi-Flu+COVID19 Ag using nasopharyngeal and anterior nasal samples, with a real-time reverse transcription PCR (RT-PCR) as a reference testing. In addition, we examined for agreement among the results of QuickNavi-Flu+COVID19 Ag, QuickNavi-COVID19 Ag and QuickNavi-Flu2.

This study was conducted at a PCR center in Tsukuba Medical Center Hospital (TMCH) located at Ibaraki prefecture, Japan between August 2, 2021 and September 13, 2021. During this study period, Japan was affected by the fifth wave of SARS-CoV-2 outbreak mainly caused by a delta strain [5]. Nasopharyngeal samples and clinical data were obtained from all patients suspected of COVID-19 and the samples were tested with an in-house PCR. In addition, we collected a nasopharyngeal sample and/or an anterior nasal sample for the evaluation of QuickNavi-Flu+COVID19 Ag with verbal informed consent. We excluded those who lacked clinical data. This study was approved by the ethics board of the University of Tsukuba Hospital Research Ethics Review Committee (approval number: R03-042).

First, we collected anterior nasal sample according to the manufacturer’s instructions, namely, inserting a swab to 2 cm in depth to one nasal cavity, rotating it five times, and holding it in place for 5 seconds. After that, nasopharyngeal samples were collected by a recommended procedure [6]. Some anterior nasal samples were collected from patients who had already been confirmed to be positive for SARS-CoV-2 on in-house PCR. These patients re-visited the hospital for checking their general conditions after their samples were confirmed SARS-CoV-2 positive. All the samples were collected with FLOQSwab (Copan Italia S.p.A., Brescia, Italy).

All antigen tests were performed on site. The swab samples were soaked in the suspension buffer included in QuickNavi-Flu+COVID19 Ag kit. From the same prepared sample, three drops were added to the device in the order of QuickNavi-Flu+COVID19 Ag (Lot No. 0041081), QuickNavi-COVID19 Ag (Lot No. 0971071), and QuickNavi-Flu2 (Lot No. 0559121). Although the manufacture has issued a recall on some lots of the QuickNavi-COVID19 Ag due to the concern for the increased false positive rate [7], our study did not include the relevant batches.

Nasopharyngeal samples collected for in-house PCR of SARS-CoV-2 were suspended in 3 mL of Universal Transport Medium (UTM) (Copan Italia S.p.A.), and the RNA was extracted with magLEAD 6gC (Precision System Science Co., Ltd., Chiba, Japan). After performing the in-house PCR [8], the remaining eluted RNA were stored at -80 °C and were transferred to Denka Co., Ltd. for reference real-time RT-PCR of SARS-CoV-2 and influenza A/B virus. The reference real-time RT-PCR methods for SARS-CoV-2 and influenza A/B virus were developed by the National Institute of Infectious Diseases (NIID), Japan [9,10]. In case of discrepancy between the in-house PCR and the reference real-time RT-PCR for SARS-CoV-2, re-evaluation using the Xpert Xpress SARS-CoV-2 and GeneXpert System (Cepheid, Sunnyvale, CA, USA) was performed, and the results were used as the final judgment (Supplementary Table 1).

The sensitivity, specificity, positive predictive value (PPV), and negative predictive value (NPV) of QuickNavi-Flu+COVID19 Ag were calculated for both nasopharyngeal and anterior nasal samples, separately using the results of the nasopharyngeal RT-PCR as comparator. Sensitivities stratified by cycle threshold (Ct) values based on N2 set in the reference real-time RT-PCR were also calculated for those with available data. The 95% confidence intervals (CIs) were calculated with the Clopper and Pearson method. All statistical analyses were conducted using the R 3.5.2 software program (The R Foundation, Vienna, Austria).

During the study period, 2375 samples were tested. Three nasopharyngeal samples without clinical data were excluded. Finally, 1510 nasopharyngeal samples and 862 anterior nasal samples were included in the analysis. Of them, 134 anterior nasal samples were obtained during the physical condition check-ups. In the nasopharyngeal sample group, 874 (57.9%) were asymptomatic, and in the anterior nasal sample group, 411 (47.7%) were asymptomatic.

Table 1 shows the diagnostic performance of QuickNavi-Flu+COVID19 Ag for SARS-CoV-2 detection using nasopharyngeal samples. Totally, the sensitivity was 80.9% (95%CI: 75.8-85.3), specificity was 99.8% (95%CI: 99.3-99.9), PPV was 98.7% (95%CI: 96.3-99.7) and NPV was 95.8% (95%CI: 94.5-96.8). In symptomatic cases, the sensitivity was 88.3% (95%CI: 82.5-92.7) and specificity was 100% (95%CI: 98.8-100). In asymptomatic cases, the sensitivity was 69.4% (95%CI: 59.9-77.8) and specificity was 99.6% (95%CI: 98.9-99.9). For Ct values < 20, the sensitivity was greater than 95% regardless of symptoms. For Ct values 25-29, the sensitivity was decreased to 46.2% (95%CI: 19.2-74.9) in asymptomatic cases, and for Ct values ≥ 30, the sensitivity was even decreased to 25.0% (95%CI: 7.3-52.4) in symptomatic cases. In SARS-CoV-2 detection, there were three cases that were positive for both QuickNavi-Flu+COVID19 Ag and QuickNavi-COVID19 Ag, but negative for both the in-house PCR and the reference real-time RT-PCR. One of the three cases had an opportunity to recollect nasopharyngeal samples when the subject came to the hospital accompanied by the confirmed COVID-19 family members re-visiting for checking general condition, and both QuickNavi-Flu+COVID19 Ag and in-house PCR were positive. Subsequently, the reference real-time RT-PCR was negative, but Xpert Xpress SARS-CoV-2 was positive (Ct value of 36.2 for E target and 37.9 for N2 target).

**Table 1.** SARS-CoV-2 diagnostic performance of QuickNavi-Flu+COVID19 Ag in nasopharyngeal samples

| Results of RT-PCR using nasopharyngeal samples* |  | All (N=1510) |  | Symptomatic (N=636) |  | Asymptomatic (N=874) |  |
| --- | --- | --- | --- | --- | --- | --- | --- |
|  |  | Positive | Negative | Positive | Negative | Positive | Negative |
| QuickNavi-Flu+COVID19 Ag | Positive | 228 | 3 | 151 | 0 | 77 | 3 |
|  | Negative | 54 | 1225 | 20 | 465 | 34 | 760 |
| Sensitivity (%) |  | 80.9 (75.8-85.3) |  | 88.3 (82.5-92.7) |  | 69.4 (59.9-77.8) |  |
| Specificity (%) |  | 99.8 (99.3-99.9) |  | 100 (98.8-100) |  | 99.6 (98.9-99.9) |  |
| Positive predictive value (%) |  | 98.7 (96.3-99.7) |  | 100 (96.4-100) |  | 96.3 (89.4-99.2) |  |
| Negative predictive value (%) |  | 95.8 (94.5-96.8) |  | 95.9 (93.7-97.5) |  | 95.7 (94.1-97.0) |  |
| Sensitivity stratified by Ct value (NIID-N2 set) (%) | < 20 | 98.4 (121/123) |  | 98.9 (86/87) |  | 97.2 (35/36) |  |
|  | 20-24 | 92.9 (79/85) |  | 95.8 (46/48) |  | 89.2 (33/37) |  |
|  | 25-29 | 70.0 (21/30) |  | 88.2 (15/17) |  | 46.2 (6/13) |  |
|  | ≥ 30 | 18.9 (7/37) |  | 25.0 (4/16) |  | 14.3 (3/21) |  |

\* The results of RT-PCR were based on the results of the NIID, Japan method [9], while the discordant cases with in-house PCR [8] were determined by Xpert Xpress SARS-CoV-2 and GeneXpert System.
Sensitivity, specificity, positive predictive value, and negative predictive value are provided with 95% confidence intervals.
Sensitivity stratified by Ct value is provided with the number of the denominator and the numerator.
RT-PCR, reverse-transcription PCR; Ct, cycle threshold; NIID, National Institute of Infectious Diseases.

Table 2 shows the diagnostic performance for SARS-CoV-2 detection using anterior nasal samples. In total, the sensitivity was 67.8% (95%CI: 61.4-73.8), specificity was 100% (95%CI: 99.1-100), PPV was 100% (95%CI: 96.5-100) and NPV was 89.3% (95%CI: 86.8-91.5). The sensitivity was 73.7% (95%CI: 65.3-80.9) in symptomatic cases and 60.0% (95%CI: 49.7-69.7) in asymptomatic cases. For Ct values < 20, the sensitivity was above 90% even in asymptomatic cases, but for Ct values 20-24, the sensitivity was decreased to 55.3% (95%CI: 38.3-71.4) in symptomatic cases, and for Ct values 25-29, the sensitivity was even decreased to 6.7% (95%CI: 0.2-31.9) in asymptomatic cases.

**Table 2.** SARS-CoV-2 diagnostic performance of QuickNavi-Flu+COVID19 Ag in anterior nasal samples

|  |  | All (N=862) |  | Symptomatic (N=451) |  | Asymptomatic (N=411) |  |
| --- | --- | --- | --- | --- | --- | --- | --- |
| Results of RT-PCR using nasopharyngeal samples* |  | Positive | Negative | Positive | Negative | Positive | Negative |
| QuickNavi-Flu+COVI D19 Ag | Positive | 158 | 0 | 98 | 0 | 60 | 0 |
|  | Negative | 75 | 629 | 35 | 318 | 40 | 311 |
| Sensitivity (%) |  | 67.8 (61.4-73.8) |  | 73.7 (65.3-80.9) |  | 60.0 (49.7-69.7) |  |
| Specificity (%) |  | 100 (99.1-100) |  | 100 (98.3-100) |  | 100 (98.2-100) |  |
| Positive predictive value (%) |  | 100 (96.5-100) |  | 100 (94.4-100) |  | 100 (91.2-100) |  |
| Negative predictive value (%) |  | 89.3 (86.8-91.5) |  | 90.1 (86.5-93.0) |  | 88.6 (84.8-91.7) |  |
| Sensitivity stratified by Ct value (NIID-N2 set) (%) | < 20 | 93.0 (106/114) |  | 93.2 (69/74) |  | 92.5 (37/40) |  |
|  | 20-24 | 63.1 (41/65) |  | 55.3 (21/38) |  | 74.1 (20/27) |  |
|  | 25-29 | 25.0 (6/24) |  | 55.6 (5/9) |  | 6.7 (1/15) |  |
|  | ≥ 30 | 14.3 (3/21) |  | 20.0 (2/10) |  | 9.1 (1/11) |  |

\* The results of RT-PCR were based on the results of the NIID, Japan method [9], while the discordant cases with in-house PCR [8] were determined by Xpert Xpress SARS-CoV-2 and GeneXpert System.
Sensitivity, specificity, positive predictive value, and negative predictive value are provided with 95% confidence intervals.
Sensitivity stratified by Ct value is provided with the number of the denominator and the numerator.
RT-PCR, reverse-transcription PCR; Ct, cycle threshold; NIID, National Institute of Infectious Diseases.

Overall, the results of QuickNavi-Flu+COVID19 Ag and QuickNavi-COVID19 Ag for SARS-CoV-2 detection were all consistent. For influenza viruses, one case in the nasopharyngeal group was positive for both influenza type A and B by QuickNavi-Flu2 but negative by QuickNavi-Flu+COVID19 Ag. This sample was confirmed negative by reference real-time RT-PCR.

In this prospective evaluation, QuickNavi-Flu+COVID19 Ag showed sensitivity of 80.9% in nasopharyngeal samples and 67.8 % in anterior nasal samples for the detection of SARS-CoV-2. Even though decreased sensitivity is known for anterior nasal samples when compared to nasopharyngeal samples, the sensitivity exceeds 90% with anterior nasal samples with high viral loads (Ct values < 20). In total of 2372 tests of QuickNavi-Flu+COVID-19 Ag, three cases were confirmed positive on the SARS-CoV-2 test line position but were negative for real-time RT-PCR.

QuickNavi-Flu+COVID19 Ag can similarly detect alpha, beta, gamma, delta, and kappa strains of SARS-CoV-2 on experimental analysis [11]. The study period coincided with the time when the L452R mutant strains accounted for approximately 80% in the community (Supplementary Figure 1) [5]. Therefore, although SARS-CoV-2 variants testing were not performed in this study, our results along with the analysis indicated that the antigen test sufficiently detect the circulating SARS-CoV-2 variant strains.

Decreased sensitivity with the anterior nasal samples when compared to the nasopharyngeal samples is known due to the lower viral load in the anterior nasal cavity [12]. However, anterior nasal sampling has the advantage of less pain for patients and less risk of droplet exposure for the examiner compared to nasopharyngeal sampling [13].

Three cases with positive results of the antigen test but negative results of the real-time RT-PCR were observed in nasopharyngeal samples for SARS-CoV-2 detection. False positives of antigen test could occur due to cross-reactions, interfering substances, extreme temperature and humidity [14]. Interestingly, in one of the three cases, results of the re-test were positive for both the antigen test and the two of three PCR examinations. The cause of this discrepancy may be due to the insufficient amount or suboptimal quality of collected samples used for the first test. Depending on pre-test probabilities of patients or epidemic situations in neighboring community, positive results of antigen tests should be carefully judged, and necessity of retesting with PCR test should be considered.

There are several limitations of this study. First, the performance of the antigen test in detecting influenza viruses could not be assessed because none of the viruses were detected by real-time RT-PCR during the study period. Second, frozen stored and transported samples were used to perform the reference real-time RT-PCR. However, the effect of cryopreservation for SARS-CoV-2 is not considered to be significant [15], and the discordant cases may be due to differences in the detectability of each PCR method.

In conclusion, QuickNavi-Flu+COVID19 Ag showed adequate sensitivity and sufficient specificity for SARS-CoV-2 detection using both nasopharyngeal and anterior nasal samples, especially in symptomatic patients.

## Data Availability

All data produced in the present study are available upon reasonable request to the authors.

## Acknowledgments

We thank Mrs. Yoko Ueda, Mrs. Mio Matsumoto, Mr. Masaomi Matsubayashi, Mrs. Yumiko Tanaka, Mrs. Mika Yaguchi, and the staff in the Department of Clinical Laboratory of Tsukuba Medical Center Hospital for their intensive support of this study.

## References

[1] World Health Organization. Coronavirus Disease (COVID-19) Situation Reports. 2021. https://www.who.int/emergencies/diseases/novel-coronavirus-2019/situation-reports (accessed October 18, 2021).

[2] Thein T-L, Ang LW, Young BE, Chen MI-C, Leo Y-S, Lye DCB. Differentiating coronavirus disease 2019 (COVID-19) from influenza and dengue. Sci Rep 2021;11:19713. https://doi.org/10.1038/S41598-021-99027-Z.

[3] Peeling RW, Olliaro PL, Boeras DI, Fongwen N. Scaling up COVID-19 rapid antigen tests: promises and challenges. Lancet Infect Dis 2021;21:e290–5. https://doi.org/10.1016/S1473-3099(21)00048-7.

[4] Otsuka Pharmaceutical Company Limited. Quick NaviTM - Flu+COVID-19 Ag Receives Regulatory Approval in Japan as a Combo, Rapid Diagnostic Kit for Influenza and COVID-19-Simultaneous testing for both viruses using a single test sample -| News Releases | Otsuka Pharmaceutical Co., Ltd.. 2021. https://www.otsuka.co.jp/en/company/newsreleases/2021/20210616_1.html (accessed October 18, 2021).

[5] National Institute of Infectious Diseases. Current Situation of Infection, September 1, 2021. 2021. https://www.niid.go.jp/niid/en/2019-ncov-e/10636-covid19-ab50th-en.html (accessed November 2, 2021).

[6] Marty FM, Chen K, Verrill KA. How to Obtain a Nasopharyngeal Swab Specimen. N Engl J Med 2020;382:e76. https://doi.org/10.1056/nejmvcm2010260.

[7] Denka Company Limited. Notice of Voluntary Recall of Certain Lots of the COVID-19 Rapid Antigen Test Kit (Second Notification). 2021. https://www.denka.co.jp/eng/storage/news/pdf/377/20211115_denka_quicknavi_covid19ag_en.pdf (accessed November 15, 2021).

[8] Kiyasu Y, Akashi Y, Sugiyama A, Takeuchi Y, Notake S, Naito A, et al. A Prospective Evaluation of the Analytical Performance of GENECUBE® HQ SARS-CoV-2 and GENECUBE® FLU A/B. Mol Diagnosis Ther 2021;25:495–504. https://doi.org/10.1007/s40291-021-00535-5.

[9] Shirato K, Nao N, Katano H, Takayama I, Saito S, Kato F, et al. Development of genetic diagnostic methods for detection for novel coronavirus 2019(nCoV-2019) in Japan. Jpn J Infect Dis 2020;73:304–7. https://doi.org/10.7883/yoken.JJID.2020.061.

[10] National Institute of Infectious Diseases. Manual for virus detection-influenza 4th ed. 2018. https://www.niid.go.jp/niid/images/lab-manual/influenza20190116.pdf (accessed October 28, 2021).

[11] Denka Company Limited. COVID-19 Rapid Antigen Test Kits Start of Sales in the U.S. Market from November ∼Our Business Partner Xtrava Health has obtained an EUA from the U.S.FDA∼. 2021. https://www.denka.co.jp/eng/storage/news/pdf/368/20211022_denka_covid19_xtrava_en.pdf (accessed October 27, 2021).

[12] Zhou Y, OLeary TJ. Relative sensitivity of anterior nares and nasopharyngeal swabs for initial detection of SARS-CoV-2 in ambulatory patients: Rapid review and meta-Analysis. PLoS One 2021;16. https://doi.org/10.1371/journal.pone.0254559.

[13] Takeuchi Y, Akashi Y, Kato D, Kuwahara M, Muramatsu S, Ueda A, et al. Diagnostic performance and characteristics of anterior nasal collection for the SARS-CoV-2 antigen test: a prospective study. Sci Rep 2021;11:10519. https://doi.org/10.1038/s41598-021-90026-8.

[14] Patriquin G, Davidson RJ, Hatchette TF, Head BM, Mejia E, Becker MG, et al. Generation of False-Positive SARS-CoV-2 Antigen Results with Testing Conditions outside Manufacturer Recommendations: A Scientific Approach to Pandemic Misinformation. Microbiol Spectr 2021;9. https://doi.org/10.1128/Spectrum.00683-21.

[15] Rogers AA, Baumann RE, Borillo GA, Kagan RM, Batterman HJ, Galdzicka MM, et al. Evaluation of transport media and specimen transport conditions for the detection of sars-cov-2 by use of real-time reverse transcription-PCR. J Clin Microbiol 2020;58. https://doi.org/10.1128/JCM.00708-20.

