## Supplementary Figure 1 for "Clinical Evaluation of a Combo Rapid Antigen Test QuickNavi-Flu+COVID19 Ag for Simultaneous Detection of SARS-CoV-2 and Influenza Viruses"

### Slide 1
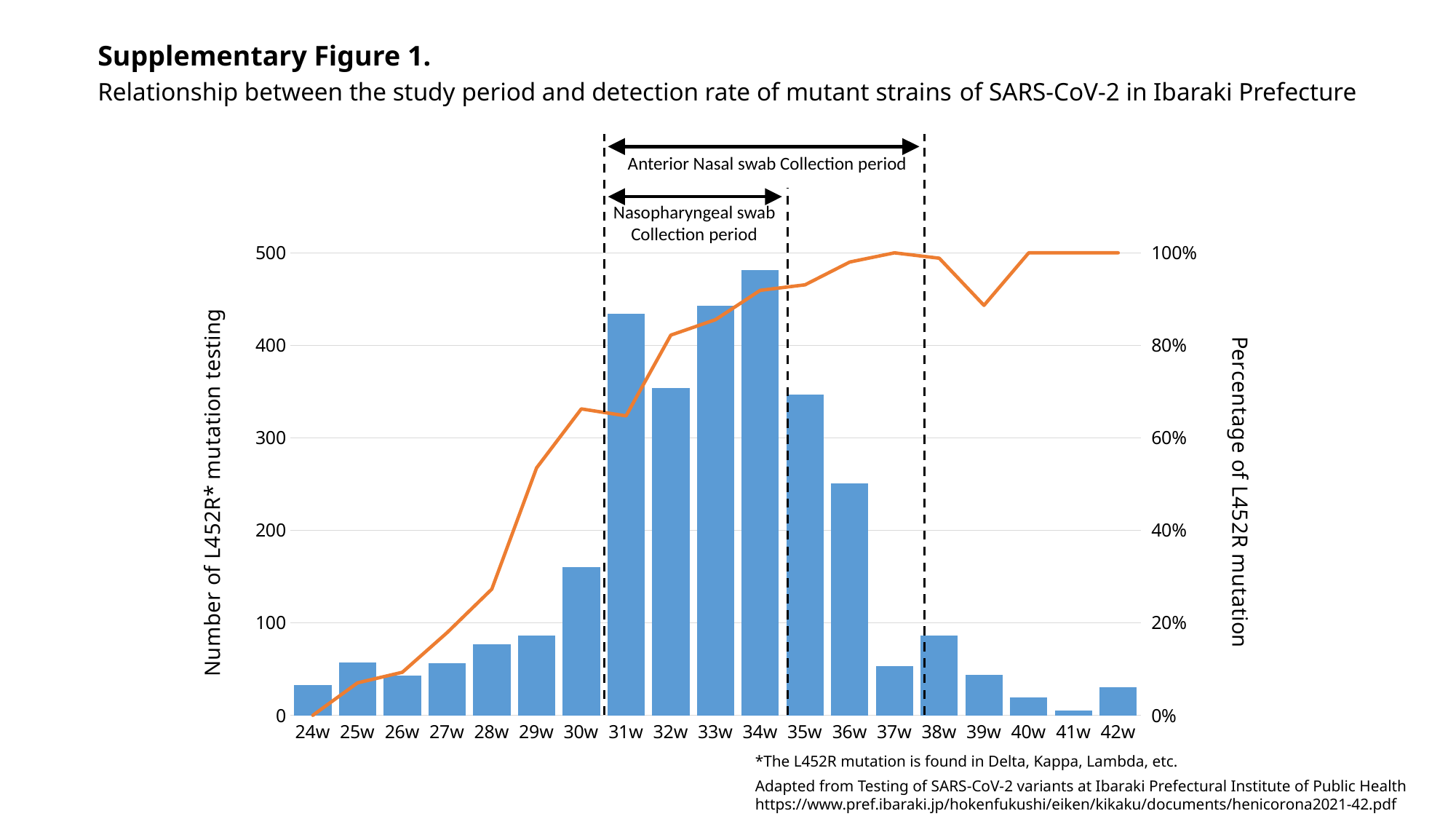

Supplementary Figure 1.
Relationship between the study period and detection rate of mutant strains of SARS-CoV-2 in Ibaraki Prefecture
Anterior Nasal swab Collection period
Nasopharyngeal swab
Collection period
#### Chart
| Category | | |
|---|---|---|
| 24w | 33.0 | 0.0 |
| 25w | 57.0 | 0.07017543859649122 |
| 26w | 43.0 | 0.09302325581395349 |
| 27w | 56.0 | 0.17857142857142858 |
| 28w | 77.0 | 0.2727272727272727 |
| 29w | 86.0 | 0.5348837209302325 |
| 30w | 160.0 | 0.6625 |
| 31w | 434.0 | 0.6474654377880185 |
| 32w | 354.0 | 0.8220338983050848 |
| 33w | 443.0 | 0.8555304740406321 |
| 34w | 481.0 | 0.918918918918919 |
| 35w | 347.0 | 0.930835734870317 |
| 36w | 251.0 | 0.9800796812749004 |
| 37w | 53.0 | 1.0 |
| 38w | 86.0 | 0.9883720930232558 |
| 39w | 44.0 | 0.8863636363636364 |
| 40w | 19.0 | 1.0 |
| 41w | 5.0 | 1.0 |
| 42w | 30.0 | 1.0 |*The L452R mutation is found in Delta, Kappa, Lambda, etc.
Adapted from Testing of SARS-CoV-2 variants at Ibaraki Prefectural Institute of Public Health
https://www.pref.ibaraki.jp/hokenfukushi/eiken/kikaku/documents/henicorona2021-42.pdf
