## Supplementary Table 1 for "Clinical Evaluation of a Combo Rapid Antigen Test QuickNavi-Flu+COVID19 Ag for Simultaneous Detection of SARS-CoV-2 and Influenza Viruses"

| Supplementary Table 1. Detailed data of 14 discordant cases between the in-house PCR and the reference real-time RT-PCR in SARS-CoV-2 detection | | | | | | | |
| --- | --- | --- | --- | --- | --- | --- | --- |
| Case No. | Symptoms | QuickNavi-Flu+COVID19 Ag | | In-house PCR | Reference real-time RT-PCR | Ct value of Xpert Xpress SARS-CoV-2* | |
|  |  | Nasopharyngeal sample | Anterior nasal sample |  |  | E target | N2 target |
| 8185 | + | Negative | Positive | Positive | Negative | 30.7 | 33.6 |
| 8738 | - | Negative | NT | Positive | Negative | 37.4 | 38.0 |
| 8921 | + | Negative | NT | Positive | Negative | 36.0 | 39.2 |
| 9192 | + | Negative | NT | Positive | Negative | 34.0 | 35.4 |
| 9343 | + | Negative | NT | Positive | Negative | UD | UD |
| 9382 | - | Negative | NT | Positive | Negative | 30.6 | 32.2 |
| 9393 | - | Negative | Negative | Positive | Negative | 31.6 | 34.0 |
| 9520 | - | Negative | Negative | Positive | Negative | 30.8 | 33.6 |
| 9703 | - | NT | Positive | Positive | Negative | 38.0 | 39.0 |
| 9707 | - | NT | Negative | Positive | Negative | 39.3 | 35.3 |
| 9832 | + | NT | Negative | Positive | Negative | 31.2 | 34.1 |
| 9856 | - | NT | Negative | Positive | Negative | 34.6 | 38.0 |
| 10186 | - | NT | Negative | Positive | Negative | 30.6 | 33.5 |
| 10267 | - | NT | Negative | Positive | Negative | 35.0 | 36.8 |
| * For analysis of this study, the Xpert Xpress SARS-CoV-2 results were used in these 14 discordant cases. | | | | | | | |
| RT-PCR, reverse-transcription PCR; Ct, cycle threshold; NT, Not tested; UD, Undetected. | | | | | | | |
